# Healthcare worker risk of COVID-19: A 20-month analysis of protective measures from vaccination and beyond

**DOI:** 10.1101/2021.12.02.21267190

**Authors:** Annalee Yassi, Stephen Barker, Karen Lockhart, Jennifer M. Grant, Arnold Ikedichi Okpani, Stacy Sprague, Muzimkhulu Zungu, Stan Lubin, Chad Kim Sing

## Abstract

**Background:** As the COVID-19 pandemic continues and new variants such as Omicron emerge, we aimed to re-evaluate vaccine effectiveness as well as impacts of rigorously implemented infection control, public health and occupational health measures in protecting healthcare workers (HCWs).

**Methods:** Following a cohort of 21,242 HCWs in Vancouver, British Columbia, Canada, for 20 months since the pandemic started, we used Cox regression and test-negative-design to examine differences in SARS-COV-2 infection rates compared to community counterparts, and within the HCW workforce, assessing the role of occupation, testing accessibility, vaccination rates, and vaccine effectiveness over time.

**Results:** Nurses, allied health professionals and medical staff in this jurisdiction had a significantly lower rate of infection compared to their age-group community counterparts, at 47.4, 41.8, and 55.3% reduction respectively; controlling for vaccine-attributable reductions, the protective impact was still substantial, at 33.4, 28.0, and 36.5% respectively. Licensed practical nurses and care aides had the highest risk of infection among HCWs, more than double that of medical staff. However, even considering differences in vaccination rates, no increase in SARS-CoV-2 infection was found compared to community rates, with combined protective measures beyond vaccination associated with a 17.7% reduced SARS-COV-2 rate in the VCH workforce overall. There was also no evidence of waning immunity within at least 200 days after second dose.

**Conclusion:** Rigorously implemented occupational health, public health and infection control measures results in a well-protected healthcare workforce with infection rates at or below rates in community counterparts. Greater accessibility of vaccination worldwide is essential; however, as implementing measures to protect this workforce globally also requires considerable health system strengthening in many jurisdictions, we caution against overly focusing on vaccination to the exclusion of other crucial elements for wider protection of HCWs, especially in facing ongoing mutations which may escape current vaccines.

*Health and care workers are the foundation of health systems and the driving force to achieving universal health coverage and global health security. …However, too many of them have become infected, ill or died as a result of COVID-19…. These deaths are a tragic loss. They are also an irreplaceable gap in the world’s pandemic response…the world cannot be complacent*. World Health Organization Steering Committee for the International Year of Health and Care Workers in 2021 (1)

## Introduction

Healthcare workers (HCWs) continue to endure much stress while the COVID-19 pandemic rages and new variants emerge, including Omicron, creating considerable new uncertainty (2). HCWs are known to be at risk of occupational exposure to Severe Acute Respiratory Syndrome coronavirus 2 (SARS-CoV-2 infection), if not adequately protected (3-9). Many experts believe that community exposure, as opposed to occupational risk factors, are the main causes of COVID-19 in HCWs (10, 11), and, in some jurisdictions, strong occupational health practices, such as ready access and well-communicated guidance on use of personal protective equipment (PPE), along with physical distancing, contact tracing and isolation requirements, have converged to keep the healthcare workforce safe at work (12). Nonetheless, the inability to “physically distance” at work in occupational groups such as flight attendants, hairdressers, and food/agriculture workers have increased risk of occupational exposure to SARS-CoV-2 infection (13-15). This is the case as well for frontline care workers in the long-term care (LTC) sector who also have shown a higher risk of SARS-CoV-2 infection (12, 16). In Canada, LTC has been disproportionately affected by the pandemic (17), and while there have been few COVID-19 deaths in HCWs across Canada, the rates of SARS-CoV-2 infection have been higher in licensed practical nurses (LPNs) and care aides, than in other HCWs (12, 18).

The World Health Organization (WHO) designated 2021 the “International Year of Health and Care Workers”, lamenting the estimated 115,500 COVID-19-related deaths among HCWs worldwide (1, 19). Additionally, while vaccination of HCWs has been a key tool in the arsenal of measures to protect the healthcare workforce (12, 20-23), the WHO noted that “available data from 119 countries suggest that by September 2021, an average of only 40% of health and care workers were fully vaccinated, with considerable difference across regions and economic groupings. Less than 1 in 10 had been fully vaccinated in the African region, while 22 mostly high-income countries reported that above 80% of their personnel were fully vaccinated.” (1). As such, the emergence of a new variant, Omicron, is particularly worrisome.

The infection prevention and control (IPAC), as well as occupational health and public health measures adopted in Vancouver, British Columbia, Canada, were described elsewhere (12). HCWs were required to wear a medical mask (ASTM level 1, 2 or 3), eye protection and gloves for all direct patient care, in addition to droplet and contact precautions when within 2 meters of COVID-19 suspect or confirmed patients. N95 or equivalent respirators were permitted based on a HCW’s point-of-care risk assessment (PCRA) and required when aerosol generating medical procedures (AGMP) were performed on a positive or suspected COVID-19 patient. Cloth and other non-approved masks were not permitted and double masking was strongly discouraged.

Physical distancing and capacity limits were created in staff common spaces and were supported by occupational health teams throughout the pandemic. IPAC measures were communicated regularly. Immunizations against COVID-19 began in December 2020, first for LTC staff, residents and essential visitors, followed by highest risk acute HCWs (Emergency room, Intensive care unit and COVID medical unit staff). Initially dose 2 was given 35 days after dose 1. The inter-dose interval was lengthened to 42 days in February 2020 then to 4 months (16 weeks) in early March 2021 to protect more people from severe disease and death (24) when vaccine supply was limited. Virtually all HCWs were vaccinated with either the Pfizer-BioNTech (93.3%) or Moderna (6.6%) COVID-19 vaccine (mRNA-1273). Vaccination of the general public started in February 2021.

In this global context, now twenty months into the pandemic and with new concerns about the sustained effectiveness of vaccines, we aimed to ascertain the ongoing effectiveness of the current package of infection control, public health and occupational health measures in a jurisdiction that has devoted considerable effort to protecting its healthcare workforce, including recently mandating full vaccination for all HCWs. As there has been considerable controversy about the effectiveness of dose schedules and vaccine program overalls, including the issue of mandating vaccination for all HCWs with its concomitant risk of losing unvaccinated HCWs from the workforce, we also aimed to explore the role of vaccination within this workforce to help tease out the extent to which COVID-19 rates in this occupational group are impacted by their higher rate of vaccination compared to the general population. Our goal in the present study was, therefore, to document SARS-CoV-2 infection rates in HCWs, ascertaining the impact of occupational role within the healthcare system on risk levels, taking community of residence and vaccination rates into account, as well as the effectiveness of the vaccination strategy to date.

## Methods

This observational cohort study was conducted among HCWs who provide services at the Vancouver Coastal Health (VCH) region, one of five health regions in British Columbia (BC), Canada. VCH provides acute, community and long-term care services to 1.25 million people which constitutes about a quarter of the population of the province. VCH is the main advanced health care referral region for the province. The VCH healthcare workforce includes 21,242 healthcare workers who provide services at 112 facilities (25).

For this study, we accessed the SARS-CoV-2 test records of all VCH HCWs who were tested between March 1, 2020, and November 11, 2021. We also obtained COVID-19 vaccination records for each HCW. Records were obtained from the provincial Workplace Health Indicator Tracking and Evaluation (WHITE™) database which is used for a variety of occupational health surveillance activities in the province. In addition to test (date and result) and vaccination (vaccine type and date received) records, we extracted details on HCW demographics and occupation including age group, sex, first 3 digits of the home residence postal code, worksite, sector, job designation, and hours worked during the period of interest. We reclassified the more than 1,000 occupational designations into seven categories: nurses, licensed practical nurses (LPNs)/care aides, administration, allied health, medical staff, support staff, others/unknown. Age groups were defined as 18-49, 50-59 and 60+. Home residence postal codes were mapped to the two local health regions: VCH and Fraser Health (FH). Data on infection and vaccination in the community were obtained from the BC Centre for Disease Control, with population projections from Statistics Canada (26). Vaccination status was categorised as unvaccinated, first dose, and second dose. We defined HCWs broadly to include anyone in any role employed/contracted by VCH who provided in-person service at VCH or in homes or community during the study period.

To avoid the possibility of reidentification of HCWs, prior to data extraction and analyses, each HCWs was assigned a unique code known only to the team statistician. No personal identifying information was included in the analytic dataset. This study was covered by the University of British Columbia Behavioural Ethics Review Board approval, certificate number H21-01138.

### Statistical analysis

Our outcome of interest was polymerase chain reaction (PCR)-confirmed SARS-CoV-2 from nucleic acid amplification test on nasal swab or gargle samples. We explored multiple exposures including occupation, home health authority (derived from postcodes), and vaccination status. We plotted the SARS-CoV-2 infection rate (per 100,000 population) and test positivity (percent of all tests performed that are positive) over time as a 7-day moving average for both HCWs and the background community from March 1, 2020, to November 11, 2021. The background community infection rates were both region and age-adjusted by weighting positive cases to match the residence and age-range distribution of the workforce. We further summarized the cumulative infection rate and vaccination status of HCWs, stratified by occupation, at the end of the observation period on November 11, 2021. To estimate vaccine effectiveness among HCWs, we first plotted the cumulative incidence of SARS-CoV-2 infection as an inverse Kaplan-Meier plot, stratified by vaccination status, starting from January 24, 2021. Then, using a Cox-proportional hazard regression model (27-30), we estimated the relative hazard ratio of one and two doses of vaccine (with unvaccinated as the reference class), adjusted for pandemic week (epi week), age group and residence. Vaccine effectiveness was calculated as 1 minus the hazard ratio determined in the Cox regression model. For comparison, we used the test-negative-design (TND) method (31-34) to estimate vaccine effectiveness, adjusting for pandemic week (epi week), age group. We explored the difference in vaccine effectiveness by the interval between the first and the second doses (less than six weeks versus more than six weeks between doses) using the TND method, and further summarised test positivity by the date of the first dose (first dose before March 1, 2021, versus on, or after, March 1, 2021). To calculate the relative risk of HCWs compared to the background community, a logistic regression model was used to determine the odds ratio. All test results from VCH and FH were included (both positive cases and negative controls from HCWs and the background community), with the region and pandemic week (epi week) of the test. We also adjusted for two-dose vaccination status; however, while vaccination status of the HCWs at the time of the test was known, the vaccination status was not published for community tests. Therefore, vaccination status for each test was included as the likelihood of vaccination (0-100%), using the region-specific two-dose community vaccination rate on the day of the test for community tests, 0% for tests on unvaccinated HCWs, 100% for tests on HCWs with two doses. Test positivity was stratified by residence and occupation, and significant differences in test positivity between strata were detected using a chi-squared test. All analyses we conducted in R version 4.1.2 (35).

## Results

Figure 1 shows the infection rate of the VCH healthcare workforce as a whole throughout the various phases of the pandemic, as well as their vaccination rates over time, along with the comparable rates in their background communities of residence, adjusting for age-differences. It could be seen that there was a spike of HCW infections at the beginning of the pandemic – likely a combination of the fact that HCWs had much better access to testing compared to the general community and consistent guidance on precautions and access to PPE. A small peak was also seen in the second wave, possibly due to outbreaks which were associated with intensive case findings efforts in HCWs, including asymptomatic testing – something not widely available for the general public. We can see that during the third wave, HCWs were well-protected, as, by this point, vaccination rates had been mounting. As for the peak in the fourth wave, this is examined in more detail below.

**Figure 1.**
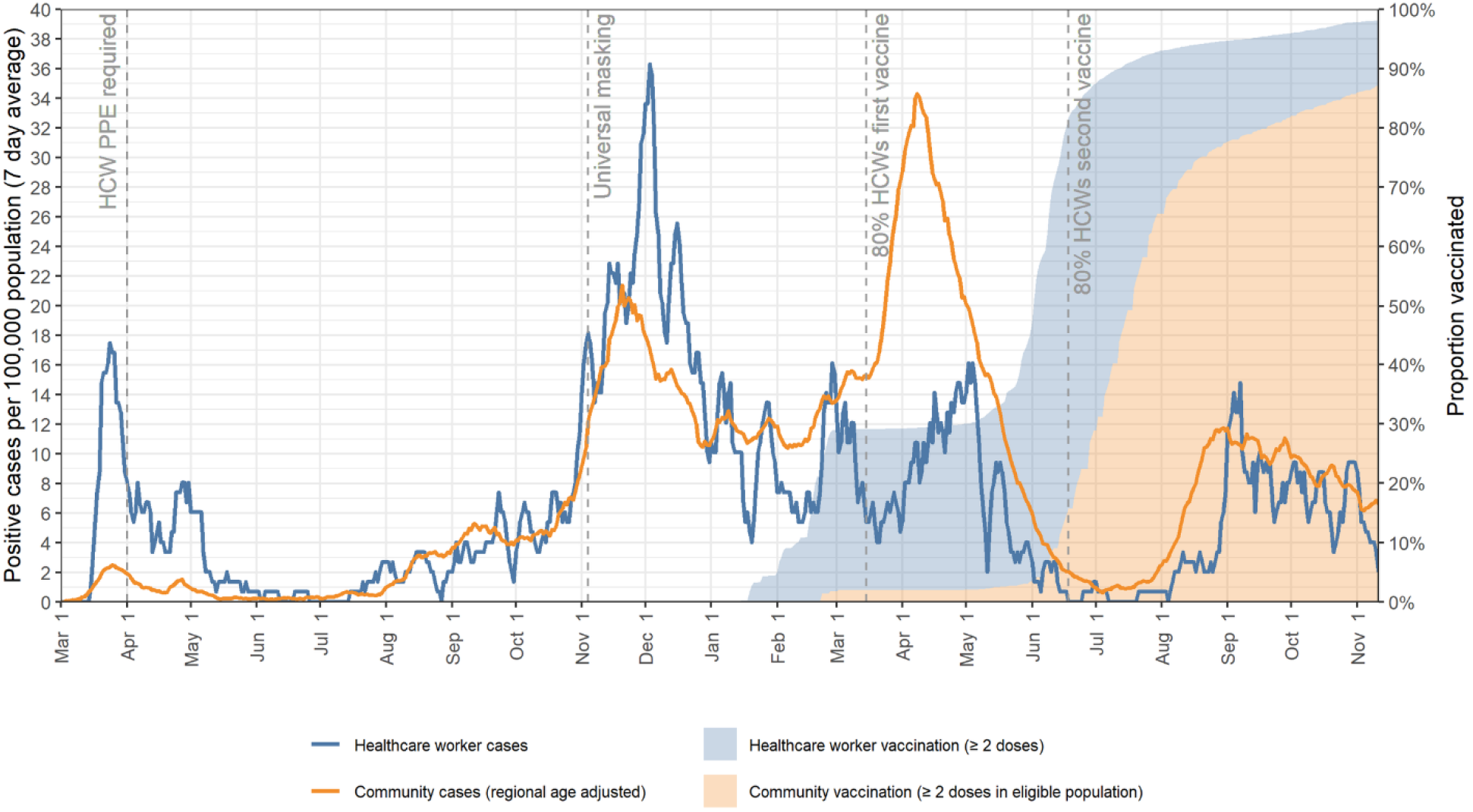
Rate of PCR-confirmed SARS-COV2 infection and vaccination rates in VCH healthcare workers and the community over time (March 1, 2020 - November 11, 2021)

Figure 2 parallels Figure 1 but shows positivity rate among those tested rather than incidence, to better account for the greater likelihood of testing among HCWs. We see that the peak that was seen in the healthcare workforce in the first wave in Figure 1, relative to the community, is no longer evident. The peak in the fourth wave, however, is more pronounced, as explored further below.

**Figure 2.**
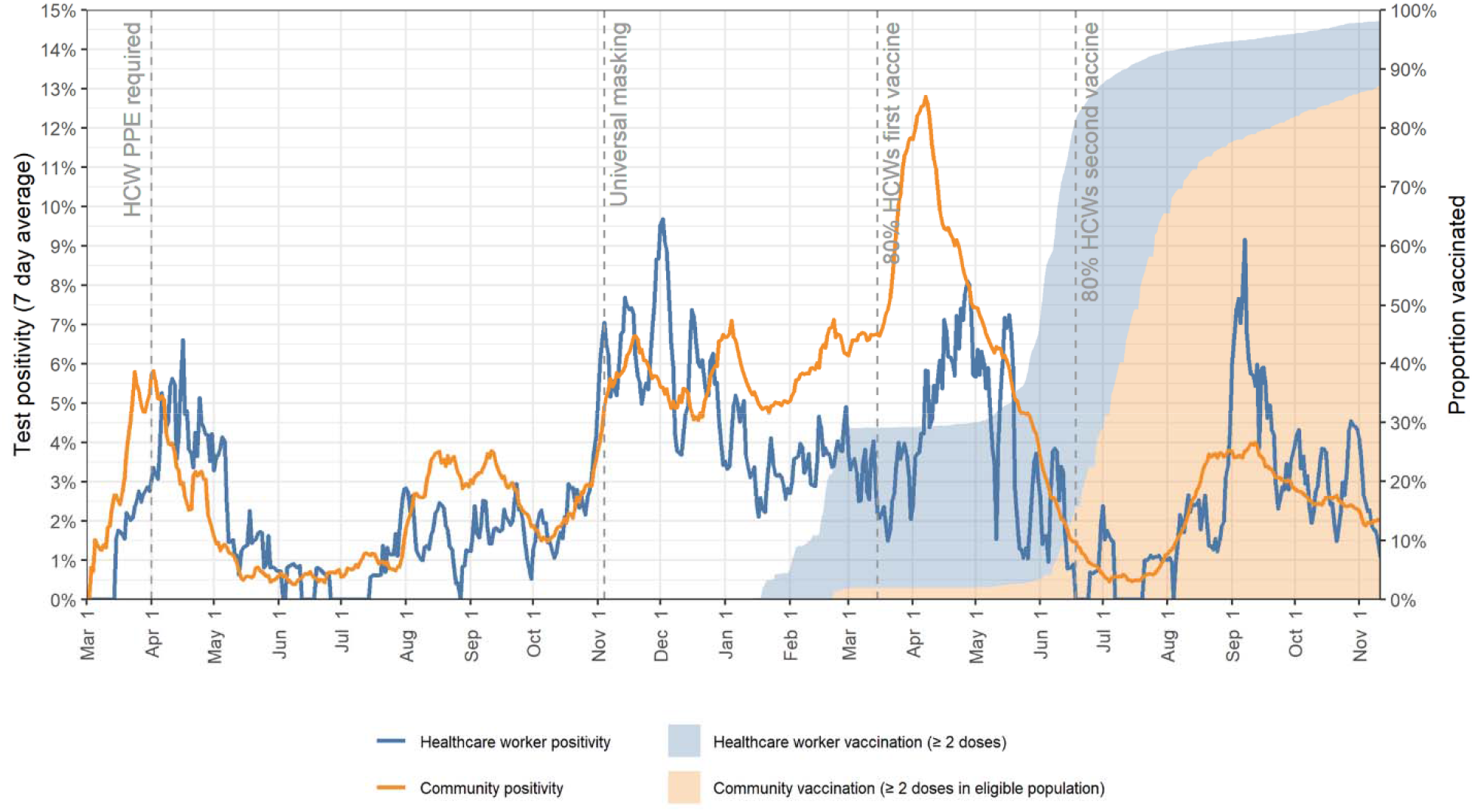
Rate of positive PCR-confirmed SARS COv2 infection amongst all VCH healthcare workers tested, and vaccination rates, along with community comparison rates, over time (March 1, 2020 - November 11, 2021)

Table 1 shows the COVID-19 overall infection rate and vaccination rate from the start of the pandemic until November 11, 2021, by occupation. It can be seen that LPNs/care aides had more than double the rate of infections as medical staff; we also see that a lower proportion of LPNs/cade aides had been vaccinated.

**Table 1.**
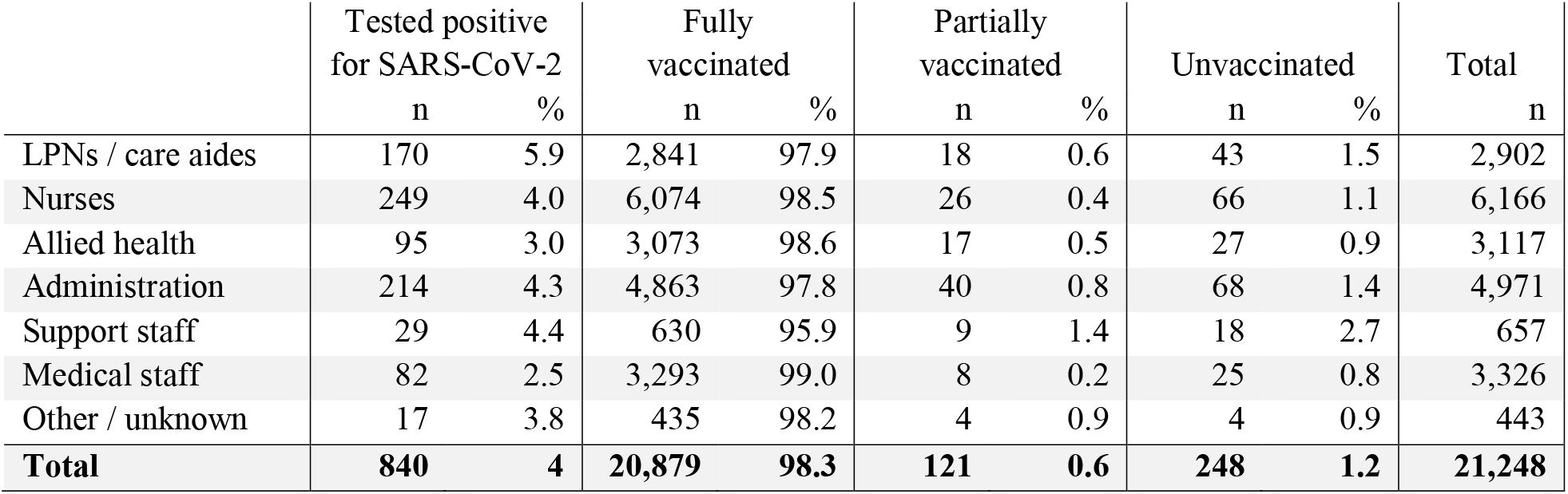
SARS-COV2 PCR-confirmed tests, and vaccination status by occupational group of VCH healthcare workers

|  | Tested positive<br>for SARS-CoV-2 |  | Fully<br>vaccinated |  | Partially<br>vaccinated |  | Unvaccinated |  | Total |
| --- | --- | --- | --- | --- | --- | --- | --- | --- | --- |
|  | n | % | n | % | n | % | n | % | n |
| LPNs / care aides | 170 | 5.9 | 2,841 | 97.9 | 18 | 0.6 | 43 | 1.5 | 2,902 |
| Nurses | 249 | 4.0 | 6,074 | 98.5 | 26 | 0.4 | 66 | 1.1 | 6,166 |
| Allied health | 95 | 3.0 | 3,073 | 98.6 | 17 | 0.5 | 27 | 0.9 | 3,117 |
| Administration | 214 | 4.3 | 4,863 | 97.8 | 40 | 0.8 | 68 | 1.4 | 4,971 |
| Support staff | 29 | 4.4 | 630 | 95.9 | 9 | 1.4 | 18 | 2.7 | 657 |
| Medical staff | 82 | 2.5 | 3,293 | 99.0 | 8 | 0.2 | 25 | 0.8 | 3,326 |
| Other / unknown | 17 | 3.8 | 435 | 98.2 | 4 | 0.9 | 4 | 0.9 | 443 |
| **Total** | **840** | **4** | **20,879** | **98.3** | **121** | **0.6** | **248** | **1.2** | **21,248** |

For analyses of the impact of vaccination on SARS-CoV-2 infection risk, we only considered the period January 24 to November 11^th^, 2021 as the numbers fully vaccinated before that date were too low to consider. First, we ascertained that of the 0.99 million unvaccinated person-days under observation within the healthcare cohort, there were 122 COVID-19 infections; for those partially vaccinated, there were 1.12 million person-days under observation, with 88 COVID-19 infections; and for the fully vaccinated 3.48 million person-days, there were 136 COVID-19 infections documented.

Using Cox regression, we found that the vaccine effectiveness (VE) of two doses, adjusted for epi-week, was 74.3% (62.8% to 82.2%). Adjusting for epi-week, age and gender, the VE was 74.1% (62.5% to 82.1%). Figure 3 shows the cumulative incidence of SARS-CoV-2 infection in the workforce by vaccine status, over time. It can be seen that HCWs who were double-vaccinated had a much lower incidence rate of SARS-CoV-2 infections, with those partially vaccinated somewhere in between unvaccinated and fully vaccinated, as would be expected. It can also be seen that the curve for those double-vaccinated does not start increasing until over 200 days post second dose, suggesting that the need for a booster dose does not appear to be urgent in this cohort; however, by 224 days post-vaccination, the curve does start increasing more sharply. Using the test-negative-design method, we considered the 6,177 tests of staff who had had 2 doses of vaccine (140 positive; 2.3% test positivity); 2,697 tests of staff with 1 dose (95 positive; 3.5% test positivity); 1,559 tests of unvaccinated staff (97 positive; 6.2% test positivity) for a total of 10,473 tests. We see the unadjusted vaccine effectiveness (VE) is 65.0% (95% CI 54.4, 73.2%). Adjusted for epi-week, we see an 82.8% (95% CI 73.9, 88.6%) rate reduction, and adjusted for epi-week, age and gender, this is still 82.8% (95% CI 74.0, 88.6%), as age and gender did not prove to be a significant covariate within our data.

**Figure 3.**
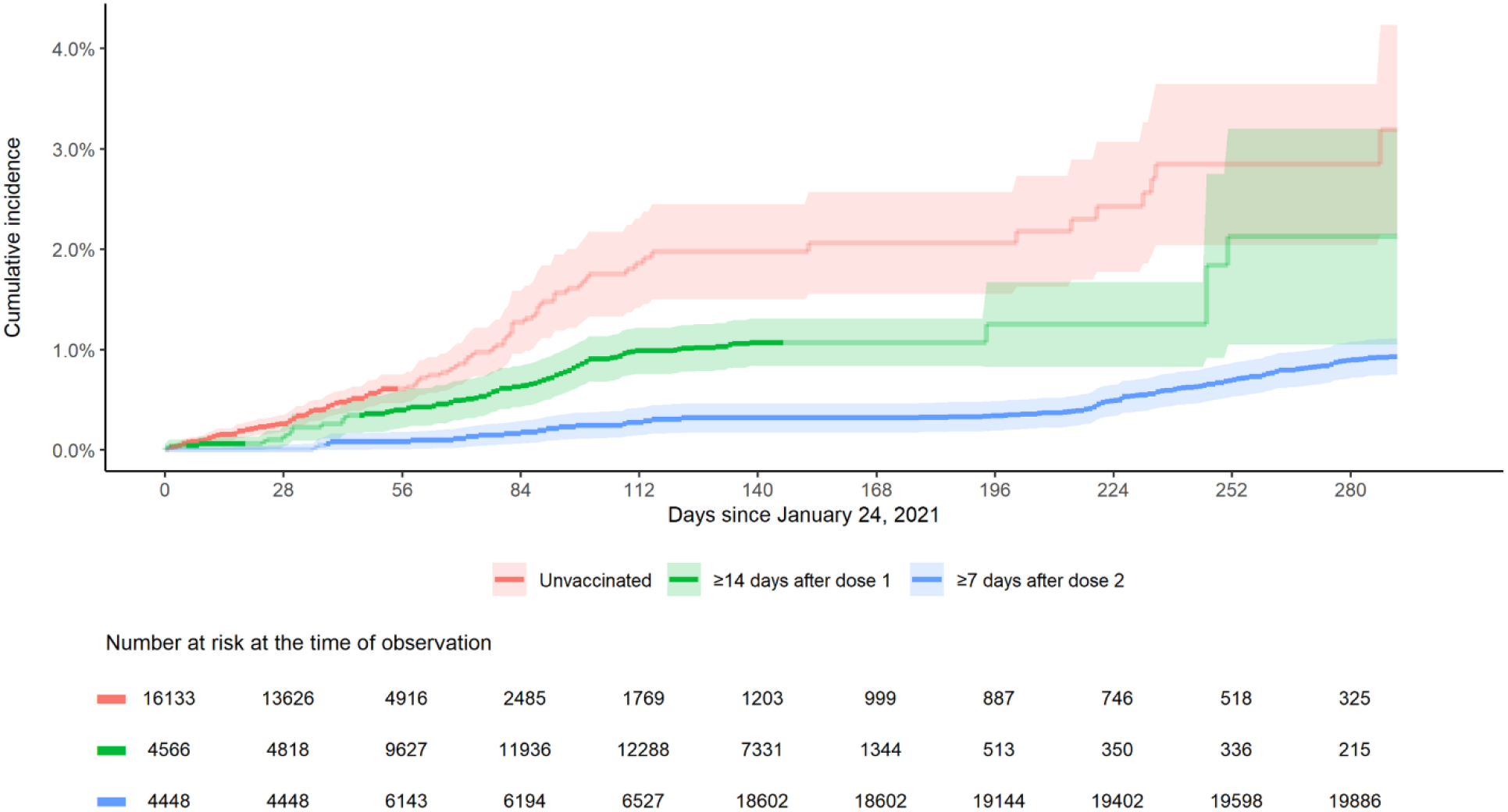
Cumulative incidence of PCR-confirmed SARS-COV2 over time in VCH healthcare workers January 24-November 11, by vaccination status

Revisiting the differing COVID-19 rates in each of the occupational groups (shown previously in Table 1) adjusting now for the differing community rates in the areas in which they live and the calendar week of the test, it can be seen from Column 2 of Table 2 that for LPNs/care aides, the rate of COVID-19 infection was 10% lower than their background community but with a wide confidence interval such that this difference is not significant. In contrast, for nurses, allied health professionals and medical staff, the lower rate compared to their communities of residence was indeed significant, with the reductions substantial, at 47.4%, 41.8%, and 55.3% respectively. When the differences in vaccination rates between HCWs and their communities of residence are taken into consideration, the difference in COVID-19 rates were 33.4%, 28.0 % and 36.5% lower, respectively. In other words, medical staff had a 55.3% reduction in their risk of COVID-19 compared to their community COVID-19 rates; further adjusting for vaccine uptake, medical staff showed a 36.5% reduction in risk of COVID-19 compared to others in their communities.Overall, we see that HCWs have lower odds of testing positive than their community counterparts, with 35.4% (95% CI 27.8, 42.3%) lower risk for HCWs when controlling for calendar time and region and 17.7% (95% CI 7.6, 26.8%) lower risk for HCWs when further controlling for vaccine uptake.

**Table 2.** COVID-19 risk reduction by occupational group within VCH healthcare workers compared to community of residence and adjusted by vaccine status

| Occupation<br>(n = number of<br>PCR tests) | Adjusting for epi-week<br>and health region of residence |  |  | Adjusting for epi-week, health region of<br>residence and vaccine status |  |  |
| --- | --- | --- | --- | --- | --- | --- |
|  | 1 – Odds<br>Ratio | 95% CI | P value | 1 – Odds<br>Ratio | 95% CI | P value |
| LPNs / care<br>aides | 10.7% | -11.4%, 28.3% | 0.317 | -24.5% | -56.1%, 0.7% | 0.057 |
| Nurses | 47.4% | 35.3%, 57.2% | <0.001 | 33.4% | 17.9%, 46.0% | <0.001 |
| Allied health | 41.8% | 21.6%, 56.8% | <0.001 | 28.0% | 2.7%, 46.7% | 0.032 |
| Administration | 13.3% | -12.2%, 33.1% | 0.278 | 2.1% | -26.9%, 24.6% | 0.87 |
| Support staff | 8.9% | -86.2%, 55.4% | 0.798 | -1.0% | -107.1%, 50.7% | 0.979 |
| Medical staff | 55.3% | 35.0%, 69.3% | <0.001 | 36.5% | 7.2%, 56.6% | 0.019 |
| Overall | 35.4% | 27.8%, 42.3% | <0.001 | 17.7% | 7.6%, 26.8% | <0.001 |

Finally, we returned to examine the COVID-19 peak in the 4th wave in HCWs to see what might account for this. We found that in the 9 weeks beginning August 29, 2021 (epi weeks 35-43), as shown in Table 3, we saw a higher test positivity rate in HCWs than the community, as 2,890 HCWs had been tested, with 106 testing positive (3.7%) over this period, while 581,385 tests were done on their community counterparts with 17,030 positive cases (2.9%). We found that the majority (97.7%) of tests performed on HCWs were on fully vaccinated HCWs (at least 1 week after the second vaccine), so it was no surprise that 98 of the 106 COVID-19 cases in HCWs were in those fully vaccinated (“breakthrough cases”). Those who were not fully vaccinated, however, had a positivity rate of 9.8%, which drove up the positivity rate overall. What was noteworthy in our refined analysis, however, was that, again, the group with the highest COVID-19 positivity was LPN/care aides, accounting for 31 of the cases during the period, with positivity of 7.5%. One of our hypotheses was that these individuals may have been HCWs who were vaccinated early and had a shorter interval between first and second dose. Indeed, as shown in Table 4, this seems to be the case; those who had been vaccinated early, with only 6 weeks between doses, had a significantly higher positivity rate than those vaccinated later and/or with a longer interval between doses.

**Table 3.** SARS-COV2 PCR-confirmed results for Vancouver Coastal Health community, Fraser Health community and VCH healthcare worker occupational groups during the 4^th^ wave (August 29-October 30, 2021)

| Category | Positive tests (n) | Negative tests (n) | Total tests (n) | Positivity (%) | P value |
| --- | --- | --- | --- | --- | --- |
| **Overall** |  |  |  |  |  |
| HCWs | 106 | 2,784 | 2,890 | 3.7% | 0.022 |
| Community | 17,030 | 564,355 | 581,385 | 2.9% | 0.022 |
| **HCW vaccine status** |  |  |  |  |  |
| Fully vaccinated | 98 | 2,710 | 2,808 | 3.5% | 0.009 |
| Not fully vaccinated | 8 | 76 | 82 | 9.8% | 0.009 |
| **HCW home residence** |  |  |  |  |  |
| HCWs living in VCH | 69 | 1971 | 2040 | 3.4% | 0.286 |
| HCWs living in FH | 33 | 731 | 764 | 4.3% | 0.286 |
| **HCW Occupation** |  |  |  |  |  |
| LPNs / care aides | 31 | 384 | 415 | 7.5% | < 0.001 |
| Nurses | 41 | 849 | 890 | 4.6% | 0.116 |
| Allied health | 9 | 500 | 509 | 1.8% | 0.015 |
| Administration | 11 | 378 | 389 | 2.8% | 0.389 |
| Support staff | 1 | 34 | 35 | 2.9% | 1.000 |
| Medical staff | 13 | 595 | 608 | 2.1% | 0.027 |

**Table 4.** SARS-COV2 PCR Confirmed test results in the 4^th^ wave in full-vaccinated VCH HCWS by date of first dose and dose schedule

| Dose schedule | First dose | Positive tests (n) | Negative tests (n) | Total tests (n) | Positivity (%) | P value |
| --- | --- | --- | --- | --- | --- | --- |
| **Less than 6 weeks** between doses | First dose **before** March 1 | 30 | 524 | 554 | 5.4% | 0.011 |
| **Less than 6 weeks** between doses | First dose on or **after** March 1 | 1 | 21 | 22 | 4.5% | 1.000 |
| **Over 6 weeks** between doses | First dose **before** March 1 | 48 | 1415 | 1463 | 3.3% | 0.487 |
| **Over 6 weeks** between doses | First dose on or **after** March 1 | 19 | 706 | 725 | 2.6% | 0.147 |
| **Overall** |  | 98 | 2,666 | 2,765 | 3.5% | 0.045 |

## Discussion

Throughout this pandemic, HCWs have, quite understandably, been concerned about their risks of COVID-19 given the nature of their jobs. Worldwide this certainly merits further attention, particularly as the pandemic is far from over and the risk of new variants, such as Omicron, is ever present. However, what we have shown is that in a jurisdiction that has devoted considerable effort to HCW protection, occupational risk can be kept to a minimum. In jurisdictions where research was able to be done in this regard, studies have generally linked HCW infection to community factors rather than to exposure in the occupational environment, Previous analysis of the Vancouver HCW cohort (12), as well as research from other well-resourced areas (36, 37), were in line with the analysis presented here: indeed VCH HCWs have incurred significantly lower rates of SARS-COV-2 infection than those of the same age-group in the communities in which they live.

However, we did find that LPNs and care aides, as well as support staff (which includes housekeeping, laundry, food services, maintenance, trades, porters, and drivers), had higher rates than other HCWs in our cohort. For these occupational groups, rates closely mirrored rates in the communities in which they live (12, 18) where it is widely believed that a host of socioeconomic factors converge to drive risk levels. These occupational groups are also comprised more often of migrants and ethnic minorities known to be at higher risk due to a myriad of social determinants (38). As adjusting for community rates and vaccine uptake did not result in the same level of reduction seen for higher-income HCWs suggest that greater patient contact in combination with perhaps less training and accessibility of resources, militates for even greater efforts to protect this component of the workforce.

We showed that vaccination reduced the risk for HCWs beyond reductions from occupational, infection control and other public health measures. Importantly, with a large proportion of the world not yet vaccinated, and many other pressing needs for healthcare resources and world attention (39, 40), our findings indicate that there does not appear to be an urgency for booster doses among HCWs in this well-protected setting, although the trend toward waning immunity from the vaccine does appear to be starting. It is also not yet known whether a booster is even needed at this stage to protect against severe disease, which, after all, is the ultimate reason for vaccination (41-43). Our data do, nonetheless, suggest that some portion of the infections in the fourth wave may be due to HCWs having been vaccinated in the early days of vaccine availability with closer dosing schedules, which may account for a higher rate of waning immunity (44); this finding coheres with other studies suggesting the value of having had a longer dose scheduling interval (45). In Israel, recent studies have shown that, despite high vaccine coverage and effectiveness, the incidence of SARS-CoV-2 has been increasing (46);

Israel used a schedule that had the second dose administered 21 days after first dose (47). As noted above, Canada proceeded with a longer interval between first and second dose, usually 16 weeks (48), except for high-risk HCWs who were vaccinated early and with a shorter dose interval.

While we have shown that rigorous implementation of infection control, occupational health and public health measures provide excellent protection, it is known that such measures are not fully in place worldwide. Zungu and colleagues, for example, demonstrated the need for system strengthening in hospitals across South Africa (49). Also Alhumaid et al. (50) documented that compliance with infection control in healthcare is associated with non-availability of resources, high workload and time limitation, as well as risk perception, caring for patients with history of infectious disease. Thus, it is important to stress that the lack of increased risk for HCWs in VCH should not be generalized to other jurisdictions where PPE and other infection control supplies may not be as easily accessible to all HCWs, or staff shortages result in breaches in proper protocols essential for staff and patient protection. Rather, the findings should provide reassurance that when evidence-based policies and procedures are implemented, revised as needed, with repeat educational messaging and rigorously monitored infection rates, this crucially important workforce can indeed be protected.

## Concluding remarks

As HCWs in this jurisdiction, and many others, face the choice of getting vaccinated or losing their job, some HCWs have asked whether vaccinations really makes a difference to their protection if they meticulously adhere to all other public health, occupational health and infection control guidelines. Our findings from analyzing 20 months of HCW surveillance data indeed reinforce the value of rigorous occupational health and infection control measures in protecting HCWs from occupational exposures. Nonetheless, we have also shown that the vaccination program for the VCH healthcare workforce has been very effective in lowering HCW risk of COVID-19 beyond the combined occupational health, public health and infection control measures implemented. And, of course, vaccination also protects against community exposures, and from severe disease. As such, it can be stated that while HCWs *in this jurisdiction* are not at increased risk of COVID-19 as a result of occupational exposures, vaccination is not only of theoretical importance to decrease the risk of transmission to vulnerable patients, but also is a measure that brings down risks for the workforce itself.

The issue of vaccination mandates is complex and subject to considerations outside the realm of medicine. From a practical point of view, mandatory vaccination may be impossible to implement due to HCW shortages, or because of philosophic objections or legal challenges. If implemented there needs to be avenues for *bona fide* medical exemptions for those with legitimate concerns, with adjudication by a neutral body with a worker-centred lens. Regardless of how vaccination is encouraged, it is clear that the HCW population can be well protected at work if rigorous occupational health and infection control measures are in place, and that vaccination provides additional benefit to workers both at work and in the community.

As the 2021 WHO Call to Action noted, shortages of health and care workers are exacerbated by the COVID-19 pandemic, with 66% of countries reporting health workforce shortages as the primary cause of disruption to essential health services (1). We stress that ensuring accessible programs of vaccination for HCWs is essential, and hoarding of vaccine by high-income countries or blocking more ready access of lower income countries to vaccine production is to be soundly opposed to ensure availability of vaccine for HCWs worldwide, the other measures that protect HCWs more broadly must not fall out of the spotlight as this pandemic moves to new stages. It must be kept in mind that HCWs face a myriad of risks, and as documented even before the pandemic, HCWs face considerable psychological distress associated with working conditions (51), with attention increasingly focused on the need for interventions to protect the mental health of HCWs (52, 53). Since the pandemic began, as also noted by the WHO in referring to the situation for HCWs globally, “levels of anxiety, distress, fatigue, occupational burnout, stigmatization, physical and psychological violence have all increased significantly” (1). Additionally, while SARS-COV2 is an immediate threat of occupational infectious transmission to HCWs worldwide, other occupational respiratory diseases, including tuberculosis, still account for high morbidity and mortality in HCWs on a global scale (54). Thus, global policies, public attention and resource allocation must keep this in mind.

## Data Availability

The data underlying the results presented in the study are available at http://innovation.ghrp.ubc.ca/COVID19/data-2021.11.29.xlsx

## Funding

This work was supported by The International Development Research Centre (IDRC) under grant M20□00559, “Protecting healthcare workers from COVID□19: a comparative contextualized analysis”. AY is supported as a Chair in Global Health from the Canada Research Council. The funders had no role in study design, data collection and analysis, decision to publish, or preparation of the manuscript.

## Acknowledgements

The authors wish to offer our gratitude to all the healthcare workers who have worked tirelessly throughout the pandemic. We also thank the many University of British Columbia medical students who staffed the Physician Occupational Safety and Health (POSH) service, as well as the Infection Prevention and Control, Public Health and People Safety teams who all work tirelessly to ensure that the healthcare workforce was well-protected. We thank the People Analytics team who extracted the data we needed for this research and the Vancouver Coastal Health staff who ensure excellent communications and a comprehensive database which guides operations and facilitates research.

## Institutional ethics review

The study was conducted according to the guidelines of the Declaration of Helsinki, and approved by the Institutional Review Board of the University of British Columbia (H21-01138).

## Data Availability Statement

The data underlying the results presented in the study are available from http://innovation.ghrp.ubc.ca/COVID19/data-2021.11.29.xlsx

## Conflicts of Interest

AY, JG, SS, CKS, SL and AO are staff or contractors of Vancouver Coastal Health.

AO, KL and SB are paid on grant funds.

AY is partially paid from a research chair through the Canadian government.

## Notes

### Summary of Updates

We are submitting a revision to our previous submission to include the figures within the manuscript file as opposed to the end of the document for ease of reading.

